# Impact of Medical Cannabis Treatment on Healthcare Utilization in Patients with Post-traumatic Stress Disorder: A Retrospective Cohort Study

**DOI:** 10.1101/2024.11.25.24317892

**Authors:** Mitchell L. Doucette, D. Luke Macfarlan, Mark Kasabuski, Junella Chin, Emily Fisher

## Abstract

**Introduction:** Medical cannabis is increasingly used as a therapy for managing post-traumatic stress disorder (PTSD). Patients with PTSD often have high healthcare utilization rates, particularly for acute services. This study examines the association between medical cannabis treatment and healthcare utilization among patients with PTSD.

**Methods:** We conducted a retrospective cohort study using cross-sectional data with temporal elements, derived from administrative records provided by Leafwell, among patients with PTSD. The cohort was defined based on medical cannabis use: the treated group included patients who had used medical cannabis for at least one year (returning for medical card renewal), while the untreated group consisted of cannabis-naive patients reporting no prior cannabis use. The primary outcomes were healthcare utilization within the past six months, including at least one urgent care visit, emergency department (ED) visit, or hospitalization related to their primary medical condition. We used inverse probability weighting with regression adjustment (IPWRA) to estimate the average treatment effect (ATE) of medical cannabis use on healthcare utilization, controlling for key demographics and health factors, including PTSD severity. Sensitivity analyses were conducted to assess the robustness of our findings.

**Results:** Among the 1,946 participants, the treated group (n = 1,261) had significantly lower healthcare utilization rates compared to the untreated group (n = 685). Using the doubly robust IPWRA model, medical cannabis treatment was associated with a significant 35.6% reduction in urgent care visits (coefficient = -0.024, Standard Error (SE) = 0.0117) and a 35.1% reduction in ED visits (coefficient = -0.027, SE = 0.0124). Hospitalization rates were 26.3% lower among the treated group but did not reach statistical significance. Sensitivity analyses utilizing alternative ATE estimation strategies displayed consistent reductions in urgent care and ED visits among cannabis users, though hospitalizations remained non-significant. Adjusting the IPWRA model’s tolerance levels strengthened the found associations while maintaining strong covariate balance. Fewer than 2% of the treated group reported an adverse event.

**Discussion:** These findings suggest that medical cannabis treatment among patients with PTSD may be associated with reduced utilization of urgent care and ED services. This relationship remains robust across multiple statistical models and sensitivity analyses, underscoring the potential role of medical cannabis in reducing acute healthcare needs in this population. Further longitudinal research is warranted to explore causality and assess its impact on hospitalization rates.

## 1 Introduction

In 2020, approximately 13 million Americans were living with post-traumatic stress disorder (PTSD) (National Center for PTSD, 2023). Throughout their lifetime, about 6 out of every 100 Americans are diagnosed with PTSD, with women (8%) being more likely than men (4%) to receive this diagnosis (National Center for PTSD, 2023). This potentially debilitating mental health condition can develop after experiencing or witnessing traumatic events, including acute injuries (Bryant, 2019; Pacella et al., 2013). Symptoms vary among individuals and may include disturbed sleep, nightmares, flashbacks to the traumatic event, and difficulty concentrating.

PTSD is also a costly medical condition. Research suggests that a new PTSD diagnosis represents an increase in total medical costs of over $6,000, adjusted for 2024 dollars (Marciniak et al., 2005). Part of the increase in total medical costs is attributable to individuals with PTSD often exhibiting a higher frequency of healthcare visits, driven by both mental and physical health challenges (Elhai et al., 2005; Kartha et al., 2008; Polusny et al., 2008). Studies underscore that those diagnosed with PTSD use a broad range of health services more frequently than those without the condition, reflecting both the disorder’s psychological impact and its interaction with physical health symptoms. Research by Elhai et al. (2005) demonstrates that trauma survivors with PTSD display elevated rates of healthcare use across multiple settings, including emergency care, hospitalizations, and outpatient mental health services. Kartha et al. (2008) found similar trends in a civilian primary care population, noting that individuals with PTSD experience a significantly higher incidence of hospital stays and mental health visits than those without PTSD. Other research notes that specific PTSD symptoms, such as avoidance, are potentially associated with increased rates of healthcare utilization as well (Polusny et al., 2008).

This pattern of utilization is further complicated by the common co-occurrence of PTSD with physical conditions, particularly chronic pain (Dahlby and Kerr, 2020; Goldstein et al., 2019; Jadhakhan et al., 2023; Pacella et al., 2013). Jadhakhan et al. (2023) identified that individuals with PTSD are at an elevated risk of developing chronic musculoskeletal pain within the first year following trauma. Many PTSD patients suffer from persistent pain, often stemming from traumatic injury, which heightens both physical and psychological distress and creates a cycle of increased healthcare demand. Part of this healthcare demand involves prescription opioids, as rates of both PTSD and opioid use disorder (OUD) have increased in the past decade (Dahlby and Kerr, 2020; Peck et al., 2021).

### 1.1 Medical cannabis and PTSD

PTSD qualifies for medical cannabis use in a majority of the 38 U.S. states where such treatment is permitted, and it is frequently reported by patients as a qualifying condition in states that allow cannabis solely for medical purposes (Boehnke et al., 2019, 2024, 2022; Doucette et al., 2024a). Cannabis exerts its pharmacological effects by interacting with the endocannabinoid system (ECS) in the body, particularly the CB1 receptor and its two endogenous ligands: N-arachidonoylethanolamide (AEA, also known as anandamide) and 2-arachidonoylglycerol (2-AG) (Hill and Gorzalka, 2009).

The hypothalamic-pituitary-adrenal (HPA) axis and the sympathetic nervous system may serve as key connections between the development of PTSD and potential ECS activation (Pervanidou and Chrousos, 2010). Current findings suggest that individuals exposed to trauma exhibit reduced cor-tisol levels either immediately or shortly thereafter, likely due to increased glucocorticoid receptor sensitivity. This reduction in cortisol leads to heightened arousal through increased noradrenergic transmission, which may contribute to the onset of PTSD. Research by Yehuda (2009) and Sarapas et al. (2011) have identified glucocorticoid signaling as a potential genetic marker for PTSD. The ECS appears responsive to glucocorticoid hormones, which may help regulate aspects of the stress response, specifically through the feedback mechanism that terminates HPA axis activity (Di et al., 2003; Evanson et al., 2010; Hill et al., 2010, 2011).

Studies involving a population-based cohort near the events of 9/11 revealed that PTSD is linked to lower circulating levels of 2-AG (Hill et al., 2013). Both endogenous CB1 receptor ligands, 2-AG and AEA, were associated with specific PTSD symptom clusters, particularly the retention of negative emotional memories. This suggests that cannabinoid-based therapies could be effective in managing certain PTSD symptoms.

An increasing body of literature supports the use of medical cannabis for managing PTSD, although much of the evidence comes from studies that do not utilize randomized controlled trials(Rehman et al., 2021; Pillai et al., 2022; Lynskey et al., 2024; Cahill et al., 2021; Sznitman et al., 2022; Nacasch et al., 2023; Roitman et al., 2014; Krediet et al., 2020; LaFrance et al., 2020). Numerous studies have reported reductions in PTSD symptom severity and improvements in sleep quality after initiating medical cannabis treatment, with patients experiencing only minimal adverse effects (Pillai et al., 2022; Lynskey et al., 2024; Cahill et al., 2021; Sznitman et al., 2022; Nacasch et al., 2023; Krediet et al., 2020). Additionally, some research has focused on how medical cannabis affects overall quality of life, showing promising outcomes in enhancing well-being (Pillai et al., 2022; Cahill et al., 2021). While only a few studies have specifically targeted PTSD patients in this regard, the positive effects on quality of life are notable. These findings suggest that medical cannabis may play a beneficial role in managing PTSD-related symptoms and improving patients’ daily functioning.

### 1.2 Current Contribution

This study seeks to quantify the impact of medical cannabis use on healthcare utilization among patients with PTSD, specifically examining urgent care, emergency department, and hospitalization rates. We estimate the average treatment effect of cannabis exposure under a potential-outcomes means, controlling for key demographic and health variables, including PTSD severity.

## 2 Methods

This retrospective cohort study analyzed cross-sectional administrative data with elements of temporality from a population of patients diagnosed with PTSD. Our outcomes of interest were healthcare service utilization, specifically having at least one urgent care visit, one emergency department visit, or one hospitalization due to PTSD symptoms in the past six months. The treated group for this study was patients with PTSD that used medical cannabis, defined as those who used medical cannabis in the past year. The untreated group for this study was patients with PTSD who did not used medical cannabis, defined as those who did not use medical cannabis in the past year. We used the doubly robust, inverse probability weighting with regression adjustment method to identify the average treatment effect of medical cannabis exposure on healthcare utilization.

### 2.1 Exposure and Outcomes Data

Outcome and exposure data were made available by the company Leafwell (Leafwell, 2024). Leafwell is a Telehealth company whose data has been used to research medical cannabis patients previously, examining demographic trends of the general population (Doucette et al., 2024a) as well as the pediatric population (Doucette et al., 2024b). Leafwell operates in 36 states in the US, and is advertised via digital media and internet search engines for connecting potential medical cannabis patients with physicians and facilitating the acquisition of a medical card if deemed medically appropriate (Doucette et al., 2024a). As part of this process, patients are asked to provide demographics and health status information after obtaining medical certification or re-certification. To accomplish our analysis, we examined Leafwell data from June 15 to September 15, 2024. Leafwell’s patient database data were collected through an online structured, cross-sectional questionnaire.

Our inclusion criteria for this study were patients who had PTSD and were either returning medical cannabis patients or new, cannabis näıve patients. We excluded new Leafwell patients who reported using cannabis for recreational or medical reasons prior to obtaining a medical card. Patients were also required to be 18 years or older.

For this analysis, we examined data from re-certifying patients, or those who obtained a medical card through Leafwell at least one year prior and returned for re-certification, as well as new patients. In this fashion, we established an element of temporality for our exposure group, as these individuals obtained a medical card via Leafwell, used medical cannabis for a year, and then returned to Leafwell to become re-certified for their medical card. Leafwell asks new patients about their past year cannabis use. Therefore, our untreated group was all new patients coming to Leafwell for the first time who self-reported their past year cannabis use as “No”.

Leafwell data were also used to determine our study’s outcomes. As part of the structured Leafwell questionnaire, both the treated and untreated groups were asked questions related to healthcare utilization in the past six months. Patients received the following question prompt, “In the past six months, can you tell us about any medical care you received related to your condition?” Patients were then able to respond “Yes” or “No” related to the statements, “I went to urgent care because of my condition,” “I went to the emergency room because of my condition,” and “I was admitted to the hospital because of my condition.” These binary variables were used as our three study outcomes. Additionally, we noted all of the adverse events reported by the treated group. Participants were asked, “When you take cannabis now, do you have any negative reactions or adverse effects?” For those who selected, “Yes,” they were further asked to specify the adverse event(s) that occurred, and they were asked to rate, on a scale from 0 to 10, with 10 being greatly impacted and 0 being no impact, how the specific adverse event(s) impacted their daily life. We provided the mean and standard deviation for the impact of each adverse event.

To protect patient confidentiality, only de-identified data from the LPD were provided to researchers, ensuring that no personally identifiable information was accessible. This study received an exemption from ethical review by an independent Institutional Review Board (BRANY, IRB Number: IRB00000080). Patients consented to the aggregate use of their questionnaire data in accordance with Leafwell’s terms of service.

### 2.2 Covariates

We selected a range of covariates based on their potential influence on both the outcome model (healthcare utilization) and the treatment model (medical cannabis exposure). For the outcome model, we included age (continuous) (Konnert and Wong, 2015; Tillmann et al., 2021), sex (male vs. female) (Kaur et al., 2007; Gaffey et al., 2021), and race/ethnicity (white non-Hispanic vs. all other races) (Husaini et al., 2004) as rates of healthcare utilization are known to vary by these demographics. We also wanted to capture elements of lifestyle choices that influence healthcare utilization. Thus, we controlled for smoking status and alcohol consumption. Both of these health behaviors are known to increase healthcare utilization, with PTSD patients having high rates of both smoking (Rosenblum et al., 2020; Fu et al., 2007) and alcohol misuse (Debell et al., 2014; Smith and Cottler, 2018; Back and Jones, 2018). We also controlled for health insurance status, as individuals with health insurance typically have higher healthcare utilization rates (Shami et al., 2019).

We also controlled for three indicators of health status related to patients’ PTSD. We controlled for PTSD severity using the Severity of Post-traumatic Stress Symptoms (NSESS) validated scale and stratified patients into either mild/moderate PTSD or severe/extreme PTSD (LeBeau et al., 2014). We used the Graded Chronic Pain Scale–Revised (Von Korff et al., 2020) validated scale to assess chronic pain severity, given the comorbidity of PTSD and chronic pain (Jadhakhan et al., 2023). We stratified chronic pain status into either no or mild chronic pain versus bothersome or high chronic pain. Lastly, we controlled for quality of life using the CDC HRQOL-4 (Moriarty et al., 2003). For this study, we stratified quality of life into two categories; those who reported having less than two unhealthy weeks in the past month (14 unhealthy days or less) versus those who reported having more than two unhealthy weeks in the past month (15 unhealthy days or more). All of the variables included in the outcome model were also included in the treatment model, except for health insurance status as health insurance status is independent of medical cannabis exposure.

### 2.3 Statistical Approach

The primary analysis used inverse probability weighting with regression adjustment (IPWRA) to estimate the average treatment effect (ATE) of medical cannabis use on healthcare utilization (Słoczyński et al., 2022; Wooldridge, 2007; Robins et al., 1994). The inverse probability weighting with regression adjustment (IPWRA) model relies on two primary assumptions to ensure robust estimation of the ATE. First, the model assumes unconfoundedness, meaning that all covariates affecting both treatment assignment and outcomes are adequately controlled in the weighting process. This assumption is crucial for ensuring that the treatment effect is unbiased and attributable to medical cannabis exposure rather than underlying differences in demographic or health characteristics. Second, the IPWRA model assumes that each individual has a non-zero probability of being assigned to either treatment group, known as the common support or overlap assumption. This ensures that the propensity scores for treated and untreated groups overlap sufficiently, enabling comparable treatment effect estimates. With these assumptions met, IPWRA provides doubly robust estimates, as it combines both inverse probability weighting and regression adjustment to minimize bias.

Propensity scores were calculated based on the selected covariates above, and overlap in propensity scores between the treated and untreated groups was assessed to confirm adequate common support (Stuart, 2010). Robust standard errors were used in the IPWRA model to enhance precision. Tests of overidentification were conducted to examine whether the IPW function achieved covariate balance (Imai and Ratkovic, 2014). We provided the probability of healthcare utilization, or the potential-outcomes means for each treatment condition, for the three healthcare utilization outcomes.

To test the robustness of our findings, we conducted two separate sensitivity analyses. First, we used three alternative ATE estimation methods: Propensity Score Matching (PSM) (Imai and Ratkovic, 2014; Rosenbaum and Rubin, 1983), Augmented Inverse Probability Weighting (AIPW) (Glynn and Quinn, 2010; Robins, 1999; Robins et al., 1994; Funk et al., 2011), and Inverse Probability Weighting with Machine Learning (IPW-ML) (Belloni et al., 2014). Each method provides a complementary approach, addressing different aspects of potential model misspecification or imbalance. The AIPW model augments the IPW framework with an additional correction term based on residuals, enhancing model robustness against minor violations in the outcome model. The IPW-ML model, with a penalty parameter optimized via cross-validation, introduces a machine learning approach to reduce potential overfitting in covariate selection. Lastly, PSM allows for matching patients with similar characteristics, further strengthening our assessment of treatment effects by reducing reliance on extrapolation in areas with limited overlap.

For the PSM method, we employed a nearest-neighbor matching approach with a caliper of 0.1, matching treated individuals to up to three untreated counterparts. AIPW added an augmentation term to account for any remaining confounding, while IPW-ML incorporated machine learning (LASSO) to optimize covariate balance in propensity score estimation.

For our second sensitivity analysis, we varied the tolerance levels in the IPWRA model (0.05, 0.075, and 0.1) to assess the impact of different levels of data inclusion on ATE estimates and model stability. For each tolerance setting, we evaluated covariate balance using the overidentification test, with p-values above 0.05 indicating adequate balance. Participant loss due to tolerance adjustment was documented to observe any effects of data exclusion on the estimates. All analyses were conducted in Stata version 18 using the teffects commands (StataCorp, 2023). Ethical guidelines for research involving human subjects were strictly followed, with institutional review board (IRB) approval obtained and participant consent provided.

## 3 Results

Table 1 outlines the demographic characteristics, health status, and healthcare utilization patterns between medical cannabis users (treated group) and non-users (untreated group). Out of 1,946 participants with PTSD, 1,261 (64.8%) were in the treated group, and 685 (35.2%) were untreated. The treated group had a lower proportion of males (42.6% vs. 56.0%, p *<*0.001) and a higher percentage of White non-Hispanic individuals (71.7% vs. 58.0%, p *<*0.001). Fewer non-drinkers were present in the treated group (41.2% vs. 53.0%, p *<*0.001). Additionally, the treated participants were older on average (mean age = 41.92 years) compared to the untreated group (mean age = 37.68 years, p *<*0.001). Health status measures indicated that the treated group reported fewer individuals experiencing three or more unhealthy weeks per month (80.7% vs. 51.6%, p *<*0.001) and a lower prevalence of bothersome or severe chronic pain (30.4% vs. 44.6%, p *<*0.001). The treated group reported higher levels of severe/extreme PTSD severity compared to the untreated group (76.3% vs. 34.6%, p *<*0.001). Health insurance coverage was higher among the treated group (84.9% vs. 76.2%, p *<*0.001). In terms of healthcare utilization, the treated group had lower rates of urgent care visits (4.0% vs. 8.9%, p *<*0.001), lower rates of emergency department visits (4.8% vs. 10.1%, p *<*0.001), and lower rates of hospitalization (2.5% vs. 4.8%, p *<*0.001). A total of 99 participants were lost due to missingness (5.1%).

**Table 1:** Demographics, Health Status, and Healthcare Utilization among Cohort.

|  | Treated | Untreated | Total | Test statistic | Missing |
| --- | --- | --- | --- | --- | --- |
| Demographics |  |  |  |  |  |
| Sex |  |  |  |  |  |
| Female | 717 (57.4%) | 301 (44.0%) | 1,018 (52.7%) | <0.001 | 13 |
| Male | 532 (42.6%) | 383 (56.0%) | 915 (47.3%) |  |  |
| Race/Ethnicity |  |  |  |  |  |
| Table continues on the next page... |  |  |  |  |  |

**Table 1: Demographics, Health Status, and Healthcare Utilization among Patients (continued).**
|  | Treated | Untreated | Total | Test statistic | Missing |
| --- | --- | --- | --- | --- | --- |
| All other race ethnicity | 357 (28.3%) | 288 (42.0%) | 645 (33.2%) | <0.001 | 1 |
| White non-Hispanic | 903 (71.7%) | 397 (58.0%) | 1,300 (66.8%) |  |  |
| **Current Smoking Status** |  |  |  |  |  |
| No | 944 (74.9%) | 488 (71.2%) | 1,432 (73.6%) | 0.084 |  |
| Yes | 317 (25.1%) | 197 (28.8%) | 514 (26.4%) |  |  |
| **Number of days with at least one alcoholic drink** |  |  |  |  |  |
| Less than seven days | 742 (58.8%) | 322 (47.0%) | 1,064 (54.7%) | <0.001 |  |
| Seven days | 519 (41.2%) | 363 (53.0%) | 882 (45.3%) |  |  |
| **Age, Mean (SE)** |  |  |  |  |  |
|  | 41.92 (0.365) | 37.68 (0.497) | 40.43 (13.16) | <0.001 |  |
| **Health Status** |  |  |  |  |  |
| **PTSD Severity** |  |  |  |  |  |
| Mild/Moderate | 158 (23.7%) | 796 (65.4%) | 954 (50.6%) | <0.001 | 62 |
| Severe/Extreme | 508 (76.3%) | 422 (34.6%) | 930 (49.4%) |  |  |
| **Quality of Life, in number of unhealthy weeks per month** |  |  |  |  |  |
| Two or less | 132 (19.3%) | 610 (48.4%) | 742 (38.1%) | <0.001 |  |
| Three or more | 553 (80.7%) | 651 (51.6%) | 1,204 (61.9%) |  |  |
| **Chronic Pain Severity** |  |  |  |  |  |
| No or Mild Chronic Pain | 862 (69.6%) | 377 (55.4%) | 1,239 (64.6%) | <0.001 | 27 |
| Bothersome or High Chronic Pain | 377 (30.4%) | 303 (44.6%) | 680 (35.4%) |  |  |
| **Health Insurance?** |  |  |  |  |  |
| No | 190 (15.1%) | 163 (23.8%) | 353 (18.1%) | <0.001 |  |
| Yes | 1,071 (84.9%) | 522 (76.2%) | 1,593 (81.9%) |  |  |
| **Healthcare Utilization, Past 6 Months** |  |  |  |  |  |
| **Visited Urgent Care 1 or more times** |  |  |  |  |  |
| No | 1,210 (96.0%) | 624 (91.1%) | 1,834 (94.2%) | <0.001 |  |
| Yes | 51 (4.0%) | 61 (8.9%) | 112 (5.8%) |  |  |
| **Visited Emergency Room 1 or more times** |  |  |  |  |  |
| No | 1,200 (95.2%) | 616 (89.9%) | 1,816 (93.3%) | <0.001 |  |
| Yes | 61 (4.8%) | 69 (10.1%) | 130 (6.7%) |  |  |
| **Hospitalized 1 or more times** |  |  |  |  |  |
| No | 1,229 (97.5%) | 652 (95.2%) | 1,881 (96.7%) | <0.001 |  |
| Yes | 32 (2.5%) | 33 (4.8%) | 65 (3.3%) |  |  |

**Note:** Test statistic t-test for age and $\chi^2$ for all other variables. SE defined as standard error.

Table 2 summarizes the results of the doubly robust IPWRA model estimating the average treatment effect of medical cannabis exposure on past six-month healthcare utilization. All outcome models had 1,847 observations, a loss of 99 participants due to missingness (5.1% of the study population). The analysis indicates that exposure to medical cannabis is associated with a statistically significant reduction in the probability of utilizing urgent care and emergency room services at least once in the past 6 months. Specifically, medical cannabis users had a 35.6% reduction in the likelihood of visiting urgent care facilities compared to non-users (coefficient = -0.0238, Standard Error (SE) = 0.0117). Similarly, the probability of emergency room visits was reduced by 35.1% among the treated group (coefficient = -0.0268, SE = 0.0124). Although there was a reduction in hospitalization rates for medical cannabis users (coefficient = -0.0100, SE = 0.0093), this difference did not reach statistical significance. The estimated probabilities show that medical cannabis users had lower utilization rates across all healthcare services assessed: urgent care (4.32% vs. 6.71%), emergency room visits (4.98% vs. 7.67%), and hospitalizations (2.81% vs. 3.81%), when compared to non-users.

**Table 2:**
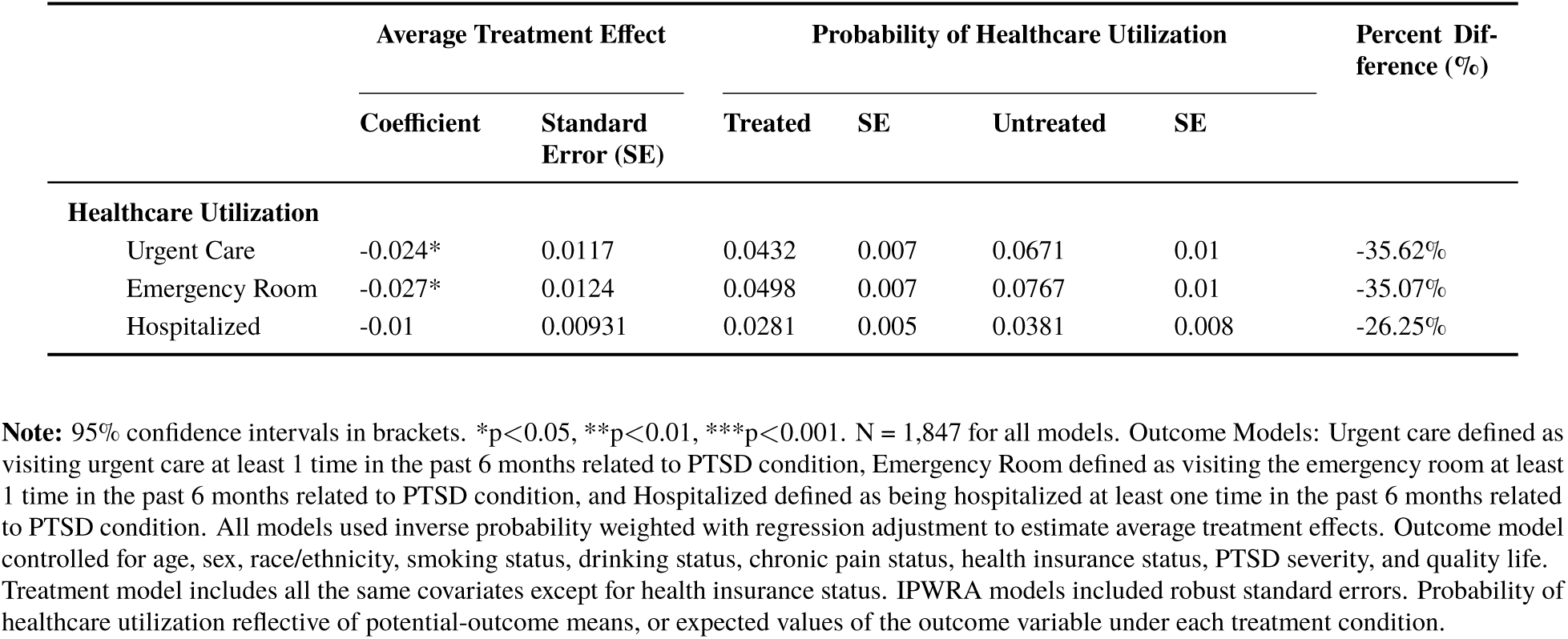
Average Treatment Effects and Estimated Probabilities of Past 6-Month Healthcare Utilization for Treated and Untreated Groups.

|  | Average Treatment Effect |  | Probability of Healthcare Utilization |  |  |  | Percent Difference (%) |
| --- | --- | --- | --- | --- | --- | --- | --- |
|  | Coefficient | Standard Error (SE) | Treated | SE | Untreated | SE |  |
| Healthcare Utilization |  |  |  |  |  |  |  |
| Urgent Care | -0.024* | 0.0117 | 0.0432 | 0.007 | 0.0671 | 0.01 | -35.62% |
| Emergency Room | -0.027* | 0.0124 | 0.0498 | 0.007 | 0.0767 | 0.01 | -35.07% |
| Hospitalized | -0.01 | 0.00931 | 0.0281 | 0.005 | 0.0381 | 0.008 | -26.25% |

**Note:** 95% confidence intervals in brackets. \* $p < 0.05$ , \*\* $p < 0.01$ , \*\*\* $p < 0.001$ . N = 1,847 for all models. Outcome Models: Urgent care defined as visiting urgent care at least 1 time in the past 6 months related to PTSD condition, Emergency Room defined as visiting the emergency room at least 1 time in the past 6 months related to PTSD condition, and Hospitalized defined as being hospitalized at least one time in the past 6 months related to PTSD condition. All models used inverse probability weighted with regression adjustment to estimate average treatment effects. Outcome model controlled for age, sex, race/ethnicity, smoking status, drinking status, chronic pain status, health insurance status, PTSD severity, and quality life. Treatment model includes all the same covariates except for health insurance status. IPWRA models included robust standard errors. Probability of healthcare utilization reflective of potential-outcome means, or expected values of the outcome variable under each treatment condition.

Figure 1 displays the overlap plot for the IPWRA. The plot illustrates the distribution of propensity scores for both the treated group and the untreated group. We observed significant overlap between the two groups across the range of propensity scores, indicating that the common support assumption is satisfied.

**Figure 1:**
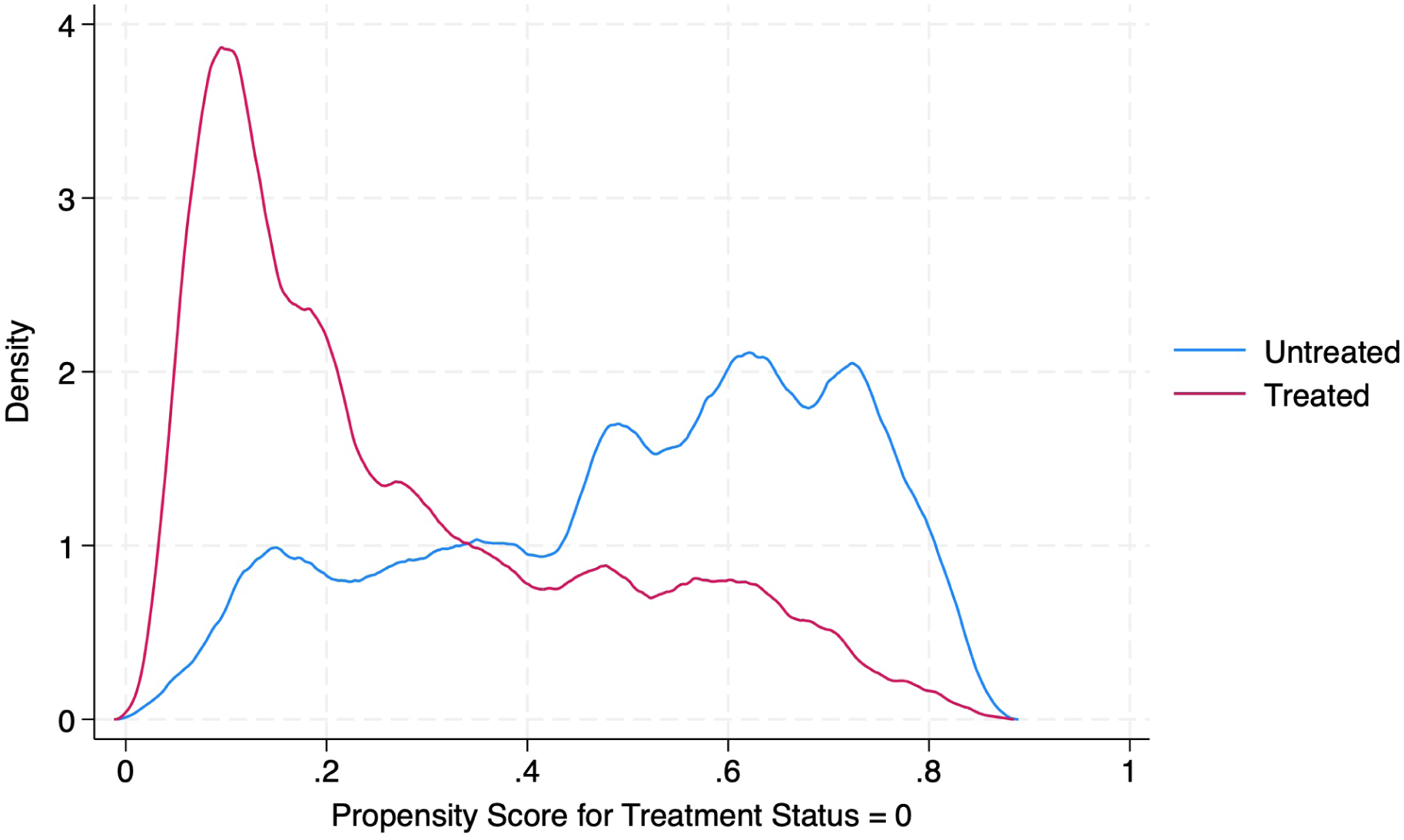
Overlap plot for Inverse Probability Weighting with Regression Adjustment.

Table 3 displays the balance diagnostics for the inverse probability weighted regression adjustment (IPWRA) model used in our analysis. The standardized differences and variance ratios for each covariate are presented both before (Raw) and after weighting (Weighted). The overall p-value of 0.212 indicates that we fail to reject the null hypothesis, suggesting that the covariates are balanced after weighting. Specifically, the standardized differences for all covariates were substantially reduced post-weighting, approaching zero, while the variance ratios moved closer to one. For example, the standardized difference for sex decreased from -0.271 to -0.028 after weighting. Similar improvements were observed for all other covariates. These results, in addition to Figure 1, suggest that the weighting procedure effectively balanced the distribution of covariates between the treated and untreated groups.

**Table 3:** Covariate balance and model diagnostics related to the inverse probability weighting with regression adjustment models.

|  |  | Standardized Differences |  | Variance Ratio |  | P-value |
| --- | --- | --- | --- | --- | --- | --- |
|  |  | Raw | Weighted | Raw | Weighted |  |
| Model Diagnostics |  |  |  |  |  |  |
| Number of observations |  | 1,847 | 1,847.00 |  |  |  |
| Treated observations |  | 1,185 | 924.6 |  |  |  |
| Untreated observations |  | 662 | 922.4 |  |  |  |
| Covariate Balance Diagnostics |  |  |  |  |  |  |
| Demographics |  |  |  |  |  |  |
| Age |  | 0.331 | -0.068 | 0.971 | 0.826 | 0.212 |
| Sex |  | -0.271 | -0.028 | 0.992 | 0.998 |  |
| Race/Ethnicity |  | 0.293 | 0.012 | 0.833 | 0.992 |  |
| Smoking Status |  | -0.058 | 0.031 | 0.940 | 1.035 |  |
| Drinking Status |  | -0.247 | 0.040 | 0.972 | 1.009 |  |
| PTSD Severity |  |  |  |  |  |  |
| Severe/Extreme PTSD |  | -0.932 | 0.001 | 1.236 | 1.000 |  |
| Quality of Life (Unhealthy Weeks) |  |  |  |  |  |  |
| Three or more unhealthy weeks per month |  | -0.648 | -0.017 | 1.600 | 1.009 |  |
| Chronic Pain Status |  |  |  |  |  |  |
| Bothersome or High Chronic Pain |  | -0.291 | -0.033 | 0.862 | 0.984 |  |

**Note:** Model diagnostics provided to display the results of the inverse probability weighting procedure. P-value results from overidentification test (*Stata* command, `teffects overid`), where the null hypothesis states that the covariates are balanced between the treatment and control groups.

We assessed the robustness of our primary findings by employing three alternative statistical methods in Table 4, PSM, AIPW, and the IPW-ML. Across all methods, the results consistently indicated that exposure to medical cannabis was associated with a reduction in healthcare utilization over the past six months, both in magnitude of association and statistical significance.

**Table 4:**
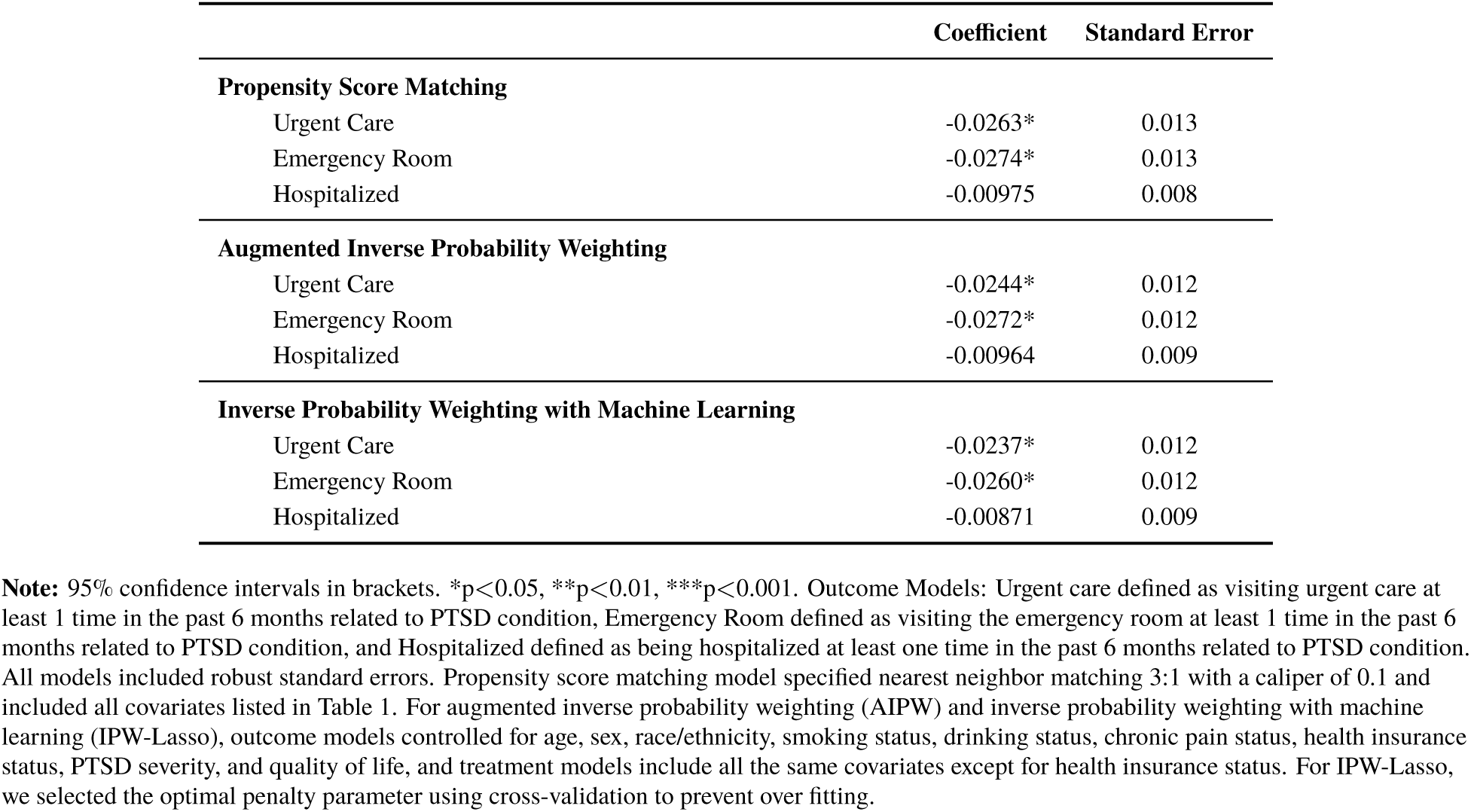
Average treatment effect of medical cannabis treatment on past 6-months healthcare utilization: Sensitivity analyses using three alternative average treatment effect estimations.

Using PSM, we found that medical cannabis patients had a statistically significant decrease in urgent care visits (coefficient = -0.0263, SE = 0.013) and emergency room visits (coefficient = - 0.0274, SE = 0.013) compared to non-patients. Although there was a reduction in hospitalization rates (coefficient = -0.00975, SE = 0.008), this difference did not reach statistical significance. The AIPW model corroborated these findings, showing significant reductions in urgent care visits (coefficient = -0.0244, SE = 0.012) and emergency room visits (coefficient = -0.0272, SE = 0.012) among medical cannabis users. Similarly, the IPW-ML approach yielded consistent results. The reductions in urgent care visits (coefficient = -0.0237, SE = 0.012) and emergency room visits (coefficient = -0.0260, SE = 0.012) remained statistically significant. Hospitalization rates did not show a significant difference in any of the models.

In our second sensitivity analysis (Table 5), we evaluated the robustness of our findings by adjusting the tolerance parameters of the doubly robust IPWRA model to 0.05, 0.075, and 0.10. At all tolerance levels, the coefficients for urgent care and emergency room visits remained statistically significant and increased in magnitude with greater tolerance. For instance, at a tolerance of 0.05, the coefficient for urgent care was -0.0245 (SE = 0.012), at a 0.075 tolerance, it was -0.0268 (SE = 0.013), and at a 0.1 tolerance, it was -0.0299 (SE = 0.013). Respectively, the percent change associated with past 6-month urgent care visits ranged from 35.6% (standard tolerance) to 38.9% (tolerance = 0.1). Similarly, the percent change associated with past 6-month emergency department visits ranged from 35.1% (standard tolerance) to 38.2% (tolerance = 0.1). The percentage of data loss increased as tolerance increased, ranging from n = 29 (1.6%) for tolerance = 0.05 to n = 255 (13.6%) for tolerance = 0.10. However, the various tolerance levels all maintained covariate balance.

**Table 5:**
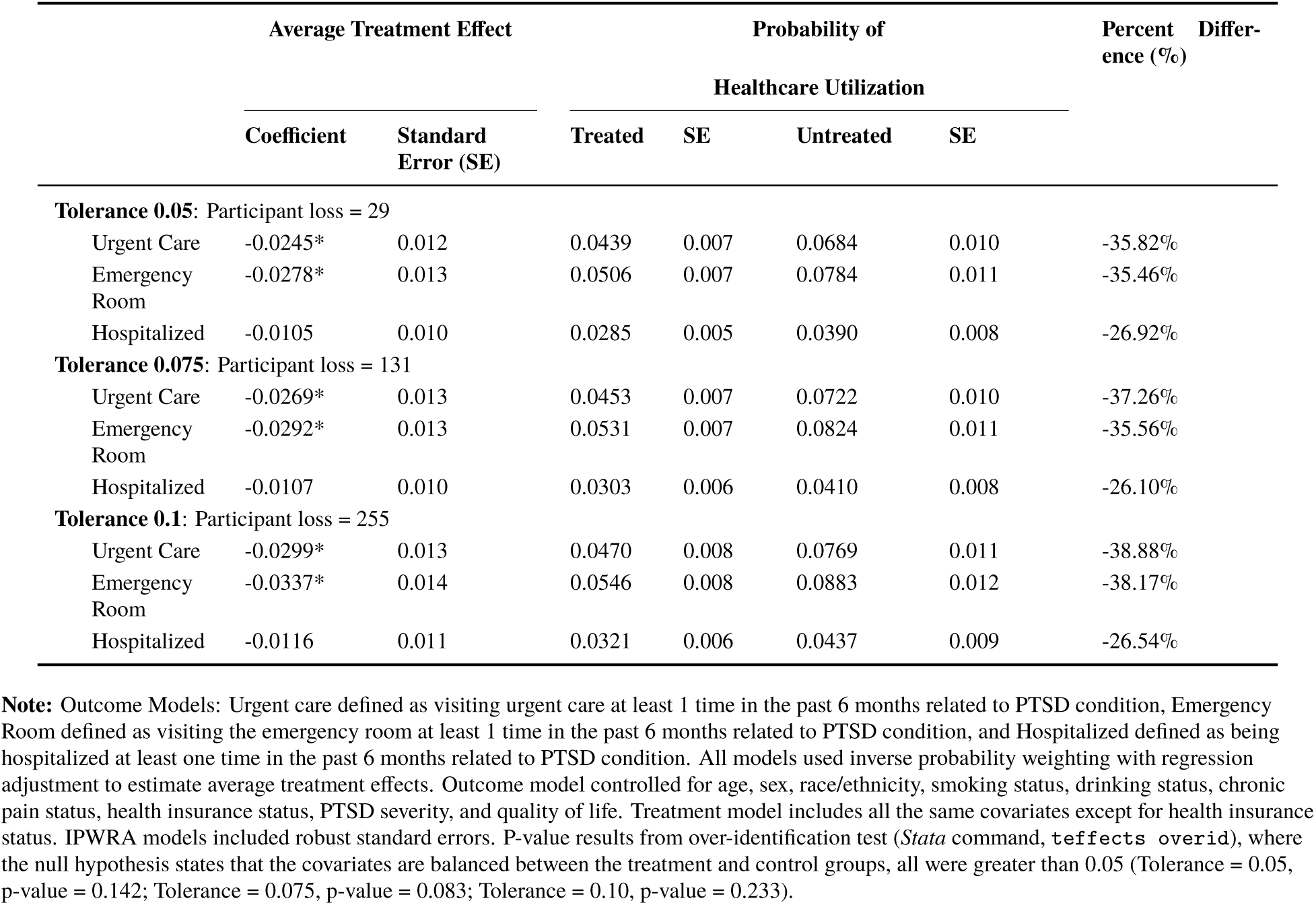
Average treatment effect of medical cannabis exposure on past 6-months healthcare utilization: Sensitivity analyses assessing the impact of different propensity score tolerance levels.

In total, 19 of the 1,949 patients with PTSD (1.51%) reported an adverse event. As participants were allowed to note one or more adverse event, there were 27 total adverse events reported. Figure 2 provides the mean and standard deviation related to the answer of, the scale of 0 to 10, “How did the adverse event impact your daily life?” We note the most common adverse events were tiredness/fatigue (n = 5) and increase in appetite (n = 5) followed by feeling sick or nauseous (n = 4). On average, participants reported that their adverse event impacted their daily life 2.43 on a scale of 0 to 10, with 10 being greatly impacted and 0 being no impact.

**Figure 2:**
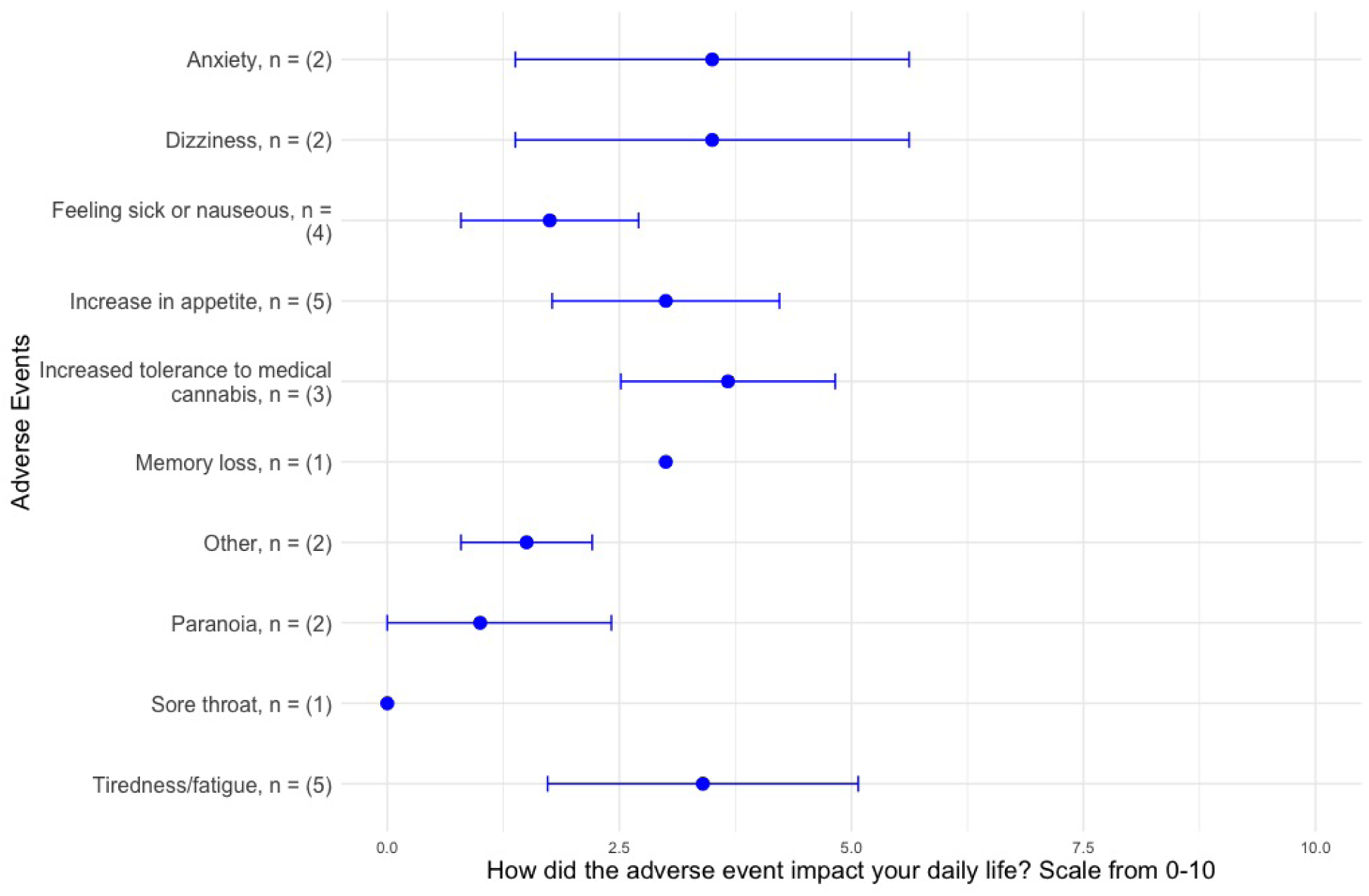
Adverse events among treated participants. The figure provides the mean and standard deviation of the impact of adverse events on participants’ daily lives on a scale from 0 to 10. The number of adverse events is provided. A total of 19 participants reported having at least one adverse event. Participants were able to report more than one adverse event if applicable.

## 4 Discussion

This study aimed to quantify the impact of medical cannabis use on healthcare utilization among patients with PTSD. Our findings indicate that exposure to medical cannabis treatment is associated with a significant reduction in urgent care and ED visits among PTSD patients. Specifically, medical cannabis users experienced a 35.6% (p *<*0.05) reduction in urgent care visits and a 35.1% (p *<*0.05) reduction in ED visits compared to non-users. Although hospitalization rates were 26.3% lower among the treated group, this difference did not reach statistical significance. The model diagnostics of our primary analysis (IPWRA) indicated that the covariates were well-balanced after weighting, with significant overlap between the treated and untreated groups, enhancing the reliability of our estimates. These results remained robust across multiple statistical models and sensitivity analyses, underscoring the potential role of medical cannabis in reducing acute healthcare needs in this population. Moreover, adverse events were minimal: only 1.5% of the treated group reported experiencing a negative reaction or adverse event, and those individuals indicated that it had a minimal impact on their daily lives.

The robustness of our findings was reinforced by the sensitivity analyses conducted. When alternative average treatment effect estimation methods—PSM, AIPW, and IPW-ML—were applied, the association between medical cannabis use and reduced urgent care and ED visits remained consistent in both magnitude and statistical significance. This consistency across various statistical techniques suggests that our results are not due to a specific modeling approach but reflect a true underlying relationship between medical cannabis use and decreased acute healthcare utilization.

Furthermore, adjusting the overlap tolerance levels in the primary IPWRA model (ranging from 0.05 to 0.1), which involved excluding observations with extreme propensity scores to improve covariate balance, showed that the reductions in urgent care and ED visits not only persisted but also increased in magnitude with stricter tolerance settings. This indicates that the observed effects are stable and not driven by outliers or specific subsets of the data. Collectively, these sensitivity analyses strengthen the credibility of our findings by demonstrating that the association is robust to different analytical methods and model specifications.

The complex interplay between trauma, the nervous system, and cannabis therapeutics reveals critical insights into managing PTSD (Pervanidou and Chrousos, 2010), with significant implications for healthcare utilization. Trauma can profoundly alter neurological functioning (Sherin and Nemeroff, 2011), triggering a persistent sympathetic nervous system response characterized by heightened anxiety, disrupted sleep, and intrusive memories. Traditional PTSD treatments often rely heavily on pharmaceutical interventions and/or psychotherapy, which can be costly (Williams et al., 2022; Burback et al., 2023). Cannabis may provide a cost-effective approach to addressing neurological dysregulation.

The observed reduction in healthcare utilization aligns with existing literature suggesting that medical cannabis may alleviate PTSD symptoms and improve patients’ quality of life (Pillai et al., 2022; Lynskey et al., 2024). Prior studies have reported that medical cannabis use leads to reductions in PTSD symptom severity, including disturbed sleep, nightmares, and flashbacks (Cahill et al., 2021; Sznitman et al., 2022). By mitigating these symptoms, medical cannabis may reduce the likelihood of acute exacerbations that necessitate urgent or emergency care. Moreover, the interaction between PTSD and chronic pain may partly explain the decreased healthcare utilization. Chronic pain is prevalent among patients with PTSD (Jadhakhan et al., 2023), and medical cannabis has been shown to have analgesic properties (Hill and Gorzalka, 2009). Chronic pain is also one of the leading qualifying medical conditions that patients cite as their reason for seeking medical cannabis (Boehnke et al., 2024; Doucette et al., 2024a). By addressing both psychological and physical symptoms, medical cannabis may offer a more comprehensive therapeutic effect, potentially reducing the need for acute healthcare services.

Our findings suggest that medical cannabis could be a valuable adjunct therapy for PTSD with minimal adverse events, potentially reducing the burden on acute healthcare services and reducing healthcare costs. Recent research suggests medical cannabis is likely a cost-effective adjunctive treatment option for moderate PTSD (Doucette et al., 2024d). Similarly, other population-level research has found medical cannabis laws are associated with reduced health insurance premiums (Doucette et al., 2024c; Cook et al., 2023). Reduced urgent care and ED visits not only benefit patients by decreasing disruptive healthcare experiences, but also alleviate strain on healthcare systems. Further research is needed to estimate the healthcare cost savings associated with our findings. Clinicians considering medical cannabis as a treatment option should weigh these potential benefits against the risks, such as drug-to-drug interactions (Graham et al., 2022).

We did not find that medical cannabis exposure was related to hospitalizations. This finding may be due to the relatively low incidence of hospitalization in our sample of close to 2,000 people. Therefore, our sample may lack the power to detect a difference. Future studies with larger samples or longer follow-up periods might clarify this relationship.

Further longitudinal research is needed to establish causality and explore the mechanisms underlying the observed reductions in healthcare utilization. Examining the dose-response relationship within the treated group, specifically, the impact of everyday versus occasional medical cannabis use on healthcare utilization among PTSD patients, could provide valuable insights. Additionally, investigating the long-term outcomes associated with varying levels of use may help identify optimal dosing regimens for effective and safe symptom management. Studies focused on different cannabis products, dosages, and modes of administration would also support the tailoring of treatments to individual patient needs. Finally, research should explore how medical cannabis interacts with other treatments, such as psychotherapy and pharmacotherapy, to optimize comprehensive care strategies for PTSD patients.

### 4.1 Limitations

A major strength of this study is the use of a large, diverse sample of PTSD patients from multiple U.S. states, enhancing the generalizability of the findings. The application of the doubly robust inverse probability weighting with regression adjustment (IPWRA) method strengthens the causal inference by controlling for a comprehensive set of covariates, including PTSD severity, comorbid chronic pain, and quality of life. However, several limitations warrant consideration. The study’s retrospective cohort design using self-reported administrative data may introduce recall bias and limit the ability to establish causality. For the treated group, there is an element of temporality, given that patients obtained a medical card and then returned at least 12 months later to re-certify with Leafwell. The exposure to medical cannabis was defined based on past-year use, without detailed information on dosage, formulation, or adherence, which could influence the outcomes. While our study controlled for many confounders, some unmeasured confounders may not have been accounted for in the models. However, our well-balanced models suggest that, after weighting, no systematic differences in covariates existed. Lastly, we examined the common support assumption by adjusting the IPWRA’s tolerance threshold for inclusion. The results showed similar ATEs to the primary analysis, with slightly larger magnitudes and more precise standard errors, suggesting our models meet the overlap assumption.

## 5 Conclusion

This study contributes to the growing body of evidence supporting the use of medical cannabis in managing PTSD symptoms. The association between medical cannabis use and reduced urgent care and ED visits highlights its potential to improve patient outcomes and reduce acute healthcare utilization. While these findings are promising, further research is necessary to fully understand the benefits, risks, and mechanisms of medical cannabis treatment in PTSD.

## Data Availability

The data used in this study are proprietary and were provided by Leafwell, a Telehealth company. Access to these data is restricted and cannot be shared publicly due to confidentiality agreements and proprietary considerations.

